# Numerical Analysis of Disastrous Effect of Reopening Too Soon in Georgia, USA

**DOI:** 10.1101/2020.07.01.20144667

**Authors:** Santanu Basu

## Abstract

Social distancing restrictions were lifted in Georgia, USA before the daily new Covid-19 cases were significantly reduced below the peak. In this paper we show through numerical analysis the disastrous consequence of this action resulting in a second peak of daily cases which caused additional fatalities.

## INTRODUCTION

Covid-19 disease caused by the new coronavirus, SARS-CoV-2 has reached almost all countries in the world by the first half of 2020. The same virus with no vaccine has caused different numbers of per capita infection and death in different countries, in different states within the same country, and even in different regions within the same state due to variation in virus management approaches. It is of great interest to quantitatively know how different disease control measures taken by the authorities have produced different results so that a set of effective measures may be developed to combat infectious diseases in the future.

The SARS-CoV-2 spreads from person to person [1]. In the most common form of transmission, the virus from an infected person’s nose or mouth enter a healthy person’s nose or mouth if the healthy person is within 6ft of the infected person without personal protection.

An effective tool used by the authorities to manage the number of casualties is social distancing measure. The idea is if any two people who usually do not live together are separated by 6 ft, then the infected person will automatically be separated by 6 ft from another healthy person who the infected person does not live together with. Another effective tool is mandating the use of face masks which prevent intake of virus by a healthy person and spread of virus by an infected person.

Since the virus stays in an infected person’s body for about 3 weeks, strict adherence to social distancing and face mask rules purges the virus from the population in low multiples of 3 weeks of residence time. A proof of this method to work is New Zealand in which the Covid-19 disease lasted for only 41 days (days with cases >5% of peak). The author has developed a Covid-19 case growth model [2–5]. The author’s model also has convincingly shown the effectiveness of this method in reducing fatalities from a new infectious disease.

Due to reasons other than protecting the health of the population, the authorities in various regions, states and countries sometimes give orders to relax social distancing measures and use of face masks which again increases the person to person virus transmission rate before the virus is substantially purged from the general population. These decisions have consequences. In this paper, we will quantitatively study the effect of premature reopening in Georgia, USA.

## OVERVIEW OF THE COVID-19 MATHEMATICAL MODEL

The author developed and documented a mathematical model to predict number of Covid-19 cases and hospital patient loads [2–5]. The model assumes that starting from a small number of infected people which act a seed, the infection grows by transmission from person to person in proximity. The model keeps track of the number of infected people at various stages of development of the disease through the course of the virus outbreak. The model is parametric and transparent so that predictions are based on parameters which are in principle measurable and the predictions can be continuously corrected as more accurate data on parameters become available. Typical machine learning and curve fitting models which are not based on any real-life parameters tend to have high degree of error, do not provide physical insight into the problem and are not suitable for carrying out what-if analyses. The present model has a number of user-supplied parameters, one of which influences the results greatly. It is the transmission factor, which by itself is time dependent and parametrized.

The objective of this paper is to analyze the number of Covid-19 cases in Georgia, USA before and after the reopening order was issued. It is hoped that comparing these two sets of data should reveal some underlying pattern which can be a valuable lesson to minimize the spread of an infectious disease and loss of lives for now and in the future.

## RESULTS AND DISCUSSION

### Before Reopening

Covid-19 case data for the state of Georgia, USA was obtained from [6]. The first case of Covid-19 in Georgia was confirmed on March 2^nd^ of 2020 and the first death was registered on March 12^th^. The authorities declared a public health state of emergency on March 16^th^, instructed people to practice social distancing on March 19^th^ and gave an order on March 26^th^ to close public schools until April 24^th^. A shelter in place order was given effective on April 3^rd^ [7].

Georgia case data until April 24^th^ could be fitted well with March 18^th^ to be the first date (n_1_) of transmission rate decrease, 14 days for exponential time constant for transmission rate decrease (G) and 5.1% for the final transmission rate (T_f_). The comparison between the model predictions and actual data on number of cases and deaths are shown in figure 1. The figures demonstrate that the model fits the actual data very well.

**Figure 1.**
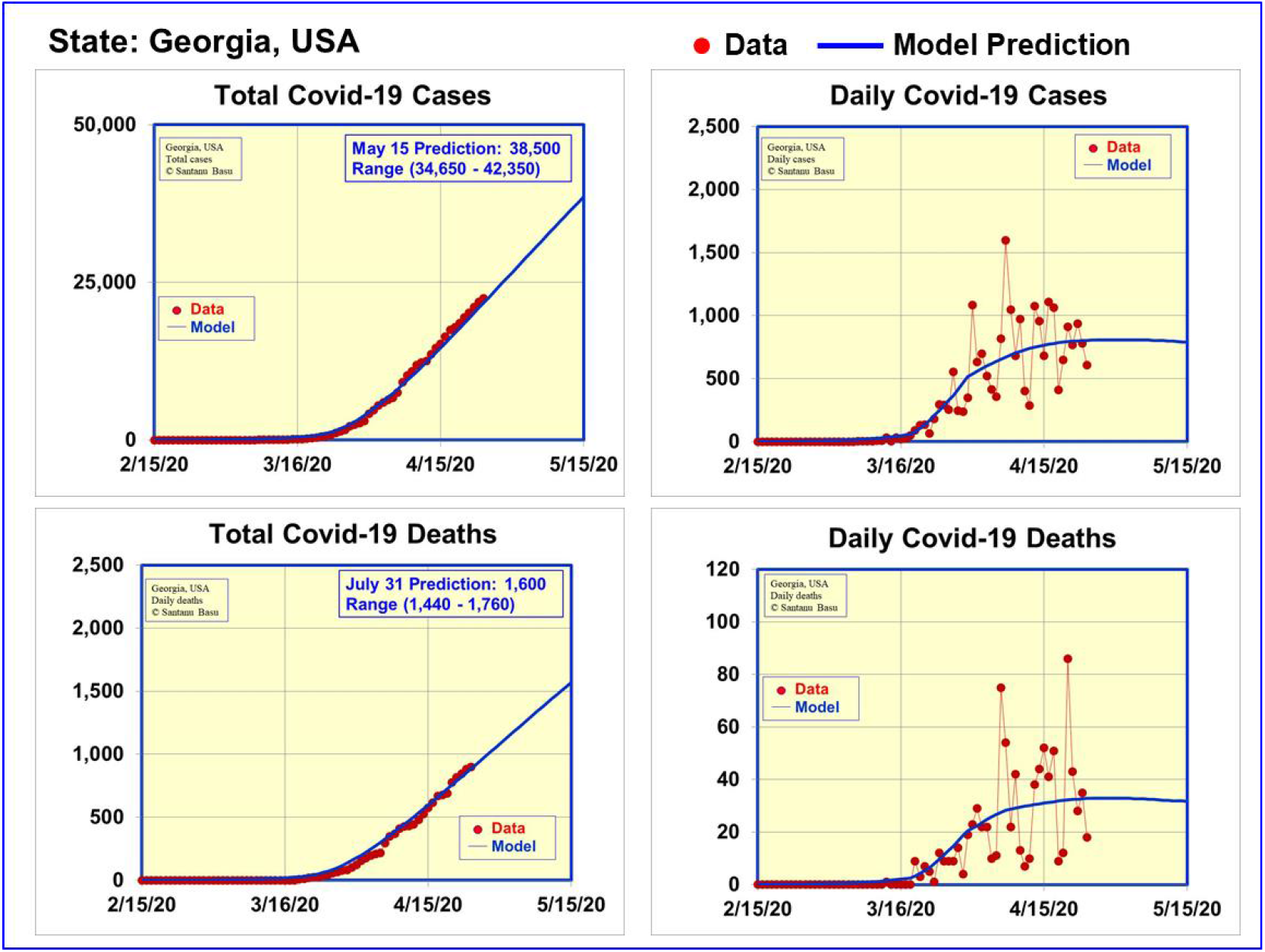
Data and model predictions for Covid-19 cases and deaths in Georgia, USA until April 24, 2020.

Figure 1 clearly shows that the positive effect of social distancing and shelter in place has not yet been realized until April 24^th^. The cumulative numbers of cases and deaths are increasing and the daily numbers of cases and deaths have not decreased. Continuing on the social distancing routine was evidently needed to being the number of infections into control.

However the same set of data as presented in figure 1 was differently interpreted by the authorities. A statement of April 20^th^ from the authorities read “According to the Department of Public Health, reports of emergency room visits for flu-like illnesses are declining, documented COVID-19 cases have flattened and appear to be declining” [7]. This seems to be at odds with the data itself.

### Reopening

The authorities gave orders to reopen businesses such as gyms and bowling alleys on April 24^th^ subject to specific restrictions, including adherence to minimum basic operations, social distancing, and regular sanitation [7].

The authorities gave orders allowing bars and nightclubs to reopen on June 1^st^ if they complied with strict sanitation and social distancing rules designed to reflect industry practices and mitigate health risk. An executive order issued on June 11^th^ stated that effective June 16^th^ residents and visitors of Georgia who are sixty-five years of age or older were no longer required to shelter in place. In restaurants and dining rooms, there was no longer a party maximum for the number of people who could sit together. There was no longer a limit on the number of patrons allowed per square foot. Workers at restaurants, dining rooms, banquet facilities, private event facilities, and private reception venues were only required to wear face coverings when they are interacting with patrons. At indoor movie theaters and cinemas, there was no longer a limit on the number of people who may sit together in a party. Walk-ins were allowed at body art studios and hair salons subject to specific requirements [7].

Figure 2 shows the disastrous consequence of the decision to reopen before the spread of infection was under control. The actual cumulative numbers of cases and deaths and daily numbers of cases and deaths between April 25^th^ and June 27^th^, 2020 are shown in figure 2. A significant rise in cases and deaths can be clearly seen that commenced on June 15^th^. The daily number of cases and deaths did not decrease at all after reopening on April 24^th^. A second peak in the daily number of cases is clearly seen in figure 2.

**Figure 2.**
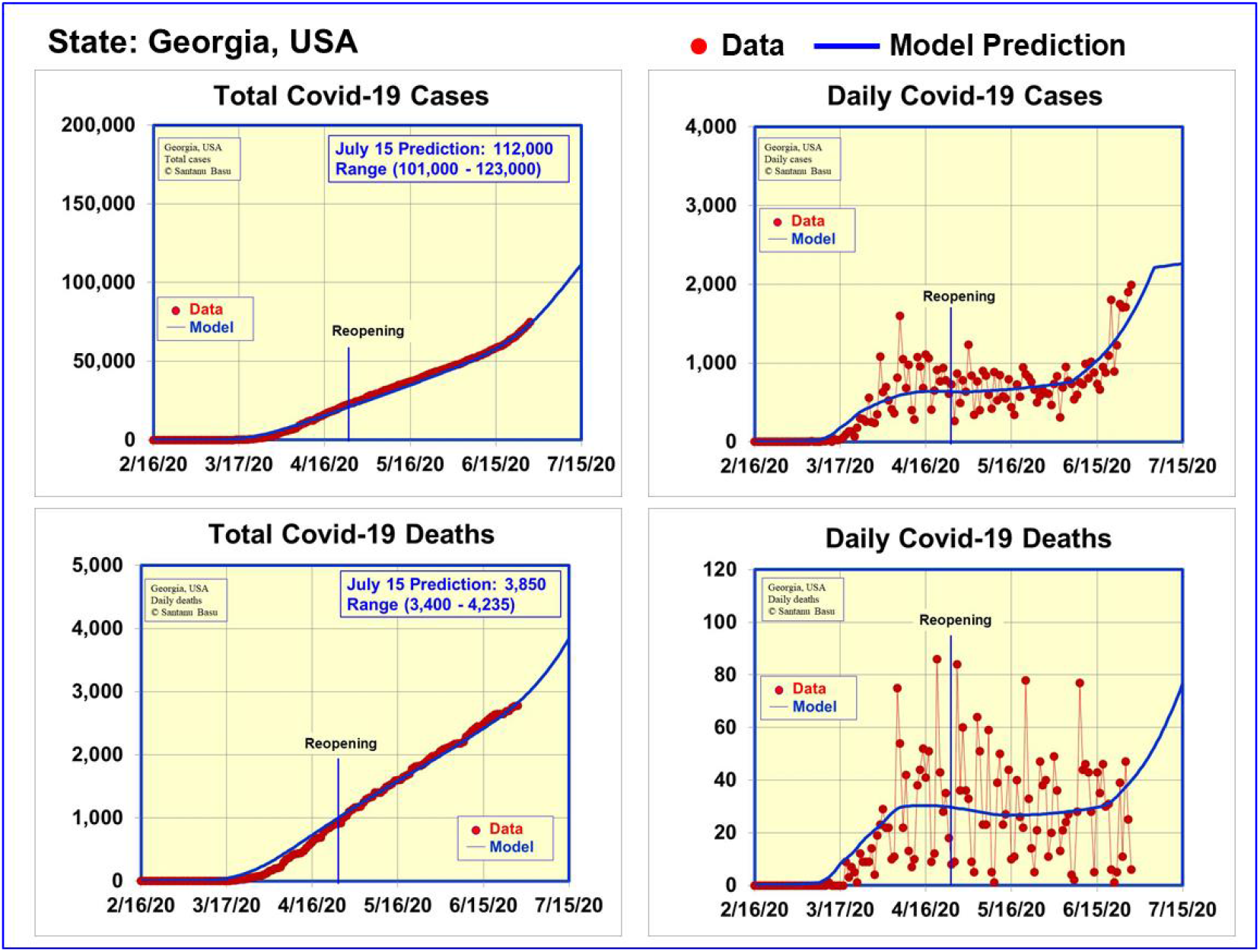
Data and model predictions for Covid-19 cases and deaths in Georgia, USA until June 27^th^, 2020.

The effect of reopening can be easily studied using our mathematical model. The reopening order led to citizens not practicing strict social distancing and wearing face masks. This in turn translated to an increase in transmission coefficient parameter of the virus from person to person. The actual time dependent transmission coefficient parameter values which we used in the model and which fit the actual case data is shown in figure 3. The transmission coefficient decreased on March 18^th^ until April 24^th^ when it started rising again as a result of reopening. The upward trend in transmission coefficient parameter accelerated starting on June 1^st^ with more of ease of restrictions.

**Figure 3.**
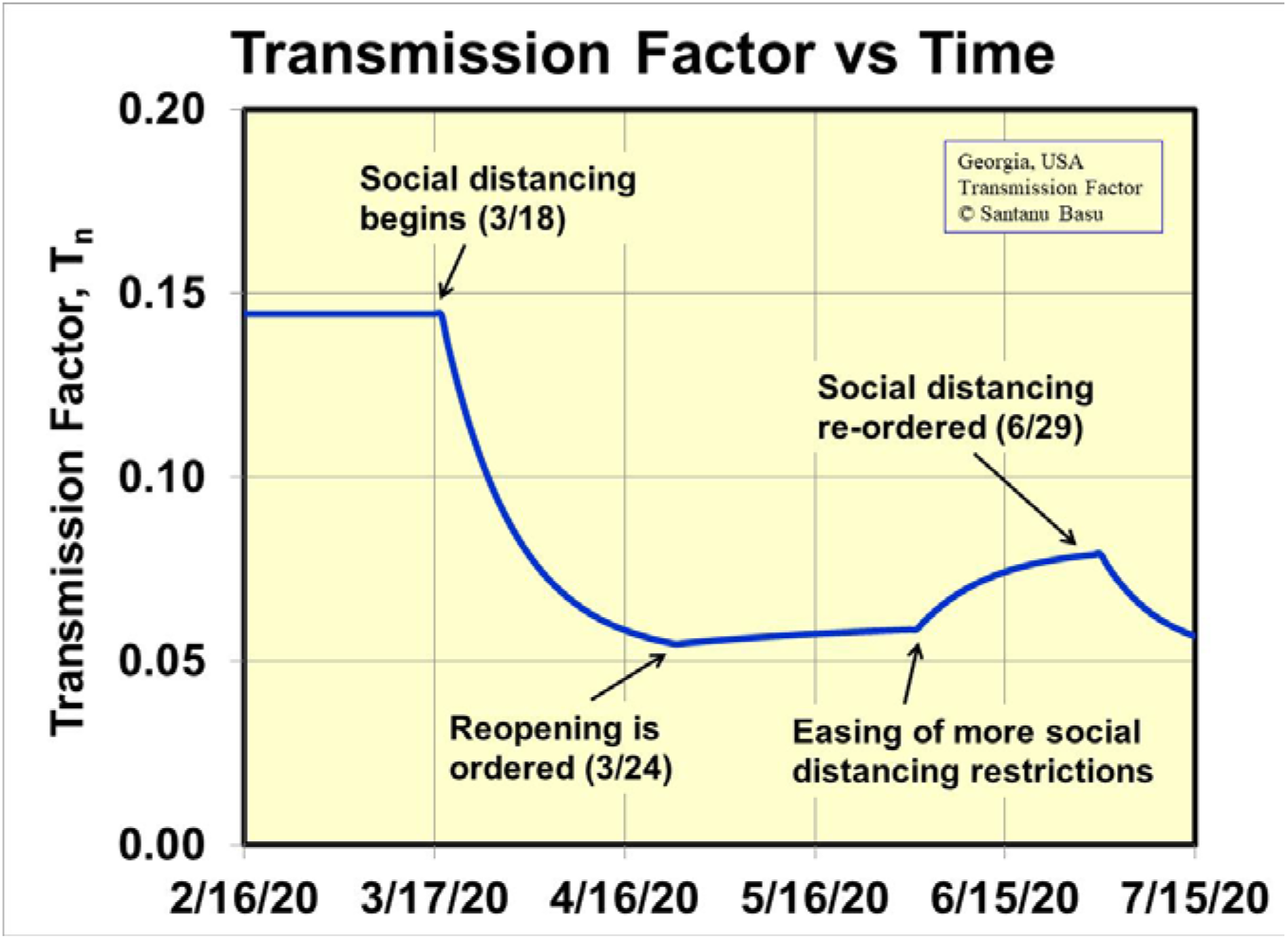
Time dependent virus transmission factor used in the mathematical model.

### Reinstatement of Social Distancing

The rise in number of Covid-19 infections prompted the authorities to issue executive orders on June 29^th^ requiring social distancing, enforcing mandatory criteria for businesses, and requiring sheltering in place for those living in long-term care facilities and the medically fragile. It is very encouraging that the authorities took steps on June 29^th^ to reinstate stricter social distancing rules [7]. Since we have a responsive mathematical model, we have updated the assumption that the transmission coefficient will begin to decrease again due to the June 29^th^ executive order. The stricter the order, the faster the transmission rate will drop and the lower will be the final transmission rate both of which will reduce casualties.

The time dependent transmission factor anchored to actual events in the state of Georgia accurately predicts the number of Covid-19 cases and deaths as shown in figure 2. This is a powerful feature of the present model.

With the model anchored to actual data until June 27^th^, we have made predictions for the expected number of cases and deaths until July 15th. The model predicts the cumulative number of cases to rise to between 101,000 and 123,000 and the cumulative number of deaths to rise to between 3,400 and 4,235. In making these predictions, we assumed that the June 29^th^ order given by the authorities would have an immediate effect on the transmission factor. The time between June 27^th^ and July 15^th^ is very critical that will determine of the trajectory of the Covid-19 cases in Georgia.

In conclusion, we have used our new Covid-19 mathematical model to quantitatively examine the consequence of premature reopening in Georgia, USA. The actual case data until April 24^th^ did not show any indication of the virus spread to be under control. It is interesting that the same data shown in figure 1 was deemed favorable by the authorities. The lesson is simple. If the reopening is done before the daily number of infections have decreased considerably, the reopening will lead to increased number of infected people and loss of lives. The necessary but difficult action of keeping people separated by about 6 ft is the key to success in this case before ultimately a vaccine becomes available.

The author wishes to acknowledge valuable discussions with P. Soni, L. Gutheinz, N. Shah, P. Hagelstein, W. Grossman, S. Gowrinathan, S. Watanabe, J. Basu, T. Hausken, S. Sheng, D. Tauber, T. Mitra, S. Dhawan, M. Genevro and J. Albertine.

## Data Availability

The data is given in the paper

https://en.wikipedia.org/wiki/COVID-19_pandemic_in_Georgia_(U.S._state)

